# Modeling suicide mortality in US counties using population socioeconomic indicators

**DOI:** 10.1101/2022.06.06.22275887

**Authors:** Sasikiran Kandula, Gonzalo Martinez-Alés, Caroline Rutherford, Catherine Gimbrone, Mark Olfson, Madelyn S. Gould, Katherine M. Keyes, Jeffrey Shaman

## Abstract

**Background:** Suicide is one of the leading causes of death in the United States and population risk prediction models can inform the type, location, and timing of public health interventions. Here, we report the development of a prediction model of suicide risk using population characteristics.

**Methods:** All suicide deaths reported to the Nation Vital Statistics System between 2005-2019 were identified, and age, sex, race, and county-of-residence of the decedents were extracted to calculate baseline risk. County-wise annual measures of socioeconomic predictors of suicide risk — unemployment, weekly wage, poverty prevalence, median household income, and population density — along with two state-wise measures of prevalence of major depressive disorder and firearm ownership were compiled from public sources. Conditional autoregressive (CAR) models, which account for spatiotemporal autocorrelation in response and predictors, were used to estimate county-level risk.

**Results:** Estimates derived from CAR models were more accurate than from models not adjusted for spatiotemporal autocorrelation. Inclusion of suicide risk/protective covariates further reduced errors. Suicide risk was estimated to increase with each standard deviation increase in firearm ownership (2.8%), prevalence of major depressive episode (1%) and unemployment (2.8%). Conversely, risk was estimated to decrease by 4.3% for each standard deviation increase in both median household income and population density. Increased heterogeneity of risk across counties was also noted.

**Conclusions:** Area-level characteristics and the CAR model structure can estimate population-level suicide risk and thus inform decisions on resource allocation and focused interventions during outbreaks.

## Introduction

Suicide rates in the United States have increased by over 30% during the last two decades, with suicide ranking among the ten most common causes of death for this period (1, 2). Along with drug overdoses and alcohol use related mortality, which have also seen large concurrent increases, suicides are responsible for the recent decrease in overall life expectancy in the US (3). Reducing suicide deaths is therefore an urgent public health challenge, and methods to predict suicide risk can be vital for determining optimal allocation of suicide prevention resources.

To date, models to predict suicide risk have largely been at the individual-level, using patient demographic characteristics and clinical history to estimate patient risk (4). Some of these models have been deployed operationally to screen patients (5), with evidence suggesting their wider adoption can be hastened through improvements in predictive ability (6-8). In contrast to individual-level models, population-level risk models have been less frequently attempted despite strong motivating factors in their favor including evidence of efficacy of population-level suicide prevention interventions, such as restrictions on access to lethal means (9-12). Population-level models can complement individual-level models as they can inform decisions on the type, location and timing of public health interventions, and provide valuable counterparts to clinical case management. In addition, when variables providing situational awareness, such as calls to crisis hotline services or posts to social media sites are also included in these models, near real-time changes in population risk can be detected, thus aiding the deployment of timely and responsive interventions. Similarly, geographically well-resolved risk estimates, can support deployment of more targeted interventions.

Risk factors for suicide have been extensively studied and include demographic characteristics such as age, race/ethnicity, sex, socioeconomic status (SES)(13-18) and mental health history (19, 20). Studies assessing the effect sizes of a combination of these characteristics, however, are relatively fewer. Meta-analyses of reported effect sizes have found considerable heterogeneity (21). Differences also exist in population-level association studies of suicide rates and risk factors (22, 23). While understanding the direction and magnitude of effects is essential, here our focus is to build on known associations to predict future suicide risk.

In this report, we describe the development of a predictive model for county level suicide risk in the US using area-level characteristics. A critical consideration when building such population-level models is the presence of spatiotemporal autocorrelation in the outcome and predictors. Inadequate accounting for this phenomenon, whereby proximate areal units during close time periods are likely to have similar observations compared to those more distant in space and time, can lead to incorrect assumptions of independence and thereby to erroneous interpretations of effects.

With deaths by suicide, spatial autocorrelation can indicate an underlying spatially correlated risk factor or a form of neighborhood effect, whereas temporal autocorrelation can be due to the same population being observed in adjacent periods, subject to the same long-term socioeconomic (SES) and environmental stressors. Furthermore, suicidal behavior has been described with contagion hypotheses and theories (24, 25). For example, acts of intentional self-harm that are directly and causally related to each other in suicide clusters, outbreaks immediately following sensationalistic media reporting of high-profile deaths by suicide (26-28), or fictional depictions of suicide (29, 30) can be seen as a contagious process (31) and lead to spatial and temporal autocorrelation (32).

Here, we model area-level risk of suicides with spatiotemporal extensions of conditional autoregressive (CAR) models, a family of Bayesian inference models commonly used in case of unmeasured spatial autocorrelation (33, 34). Comprehensive reviews of these methods are available elsewhere (35, 36). The CAR model form used in this study is an ANOVA-style decomposition of the variation in disease risk into separate sets of spatial random effects, temporal random effects and independent space-time interactions (henceforth CAR-ANOVA).

Our overall objective was to develop and evaluate the feasibility of a model for suicide risk in US counties while accounting for spatiotemporal autocorrelation in predictors and outcome. The model thus built was used to:

- Quantify the effect estimates of SES covariates on suicide risk;
- Quantify annual national suicide risk in the US and changes in heterogeneity of county-level risk during years 2005-2016; and
- Assess the accuracy in predicting yearly county-level suicide mortality risk and measure improvements relative to commonsense baseline risk estimates.

## Materials and Methods

We used a variety of public data sources for estimates of area-level suicide mortality risk factors. Detailed mortality records were obtained through a request to the National Center for Health Statistics. The explanatory variables are briefly described here and in Table 1 (see Appendix Text 1 for additional details).

- *Proportion of population living in poverty* and *median household Income:* Annual county-level measures of poverty prevalence and median household income as estimated by the US Census Bureau’s Small Area Income and Poverty Estimates (SAIPE) program (37).
- *State prevalence of major depressive episodes:* Estimates of the proportion of population in a state with at least one major depressive episode during the previous year as available in the National Surveys on Drug Use and Health (NSDUH) dataset (38). As data at county resolution were not available, prevalence was assumed to be the same in all counties in a state.
- *State prevalence of firearm-owning households:* Annual estimates of the proportion of adults who live in a household with firearms for each state in the US, provided by the RAND’s Household Firearm Ownership Database(39). Ownership rates were assumed to be the same in all counties in a state.
- *Average weekly wage:* County-level estimates of annual average weekly wage across all industries as reported to the Quarterly Census of Employment and Wages program (40) of the US Bureau of Labor.
- *Unemployment rate:* County-level estimates of unemployment estimated from standard surveys state unemployment insurance systems by the Labor and Unemployment Statistics(41) program of the US Bureau of Labor.
- *Population Density:* Annual county-level population density estimated from intercensal and postcensal population estimates and county land area (42).

**Table 1.** Variables used in the study, along with data source, description, available period and geographical resolution. Mean and standard deviations are reported for 2005-2016 for all variables.

| Variable | Data Source | Description | Available Period | Geographical resolution | Mean (SD) |
| --- | --- | --- | --- | --- | --- |
| *y* | NVSS | Observed suicide-related deaths | 2003 - 2019 | County | 12.3 (32) |
| *unemp* | BLS | Unemployment rate | 2003 - 2019 | County | .067 (.03) |
| *wage.wk* | BLS | Weekly wage, US\$ | 2003 - 2019 | County | .664 (.16) |
| *poverty.percent* | SAIPE | Proportion of population living in poverty | 2003 - 2019 | County | .162 (.06) |
| *median.hh.income* | SAIPE | Median household income, US\$ | 2003 - 2019 | County | .044 (.01) |
| *firearm.rate* | RAND | Proportion of population living in firearm-owning households | 2005 - 2016 | State | .412 (.11) |
| *mde* | NSDUH | Proportion of population with major depressive episode | 2005 - 2018 | State | .069 (.01) |
| *pop.density* | Census | Population per square mile, log | 2005 - 2019 | County | 3.77 (1.8) |

### Suicide Mortality (outcome)

Records of all-cause deaths were obtained from the US National Vital Statistics System (43). Deaths resulting from suicide were identified using *International Classification of Diseases, Tenth Revision* underlying cause-of-death codes X60-X84, Y87.0, and U03(44). County estimates for total population and population stratified by age and gender were obtained from the Bridged-Race Intercensal (2005-2009)(45) and Postcensal (2011-2019) (46) datasets and used to calculate annual, county-level suicide mortality risk.

### Conditional Autoregressive (CAR) Models

A CAR model under Poisson distribution assumption is specified as:

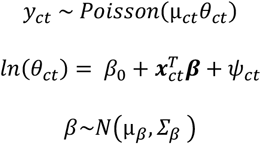

where *y*_*ct*_ denotes observed count of suicide deaths in county *c* during year *t*, µ_*ct*_ is the expected suicide deaths in county *c* in year *t* and *θ*_*ct*_ is the risk relative to µ_*ct*_ (see Appendix Text 2). ***x***_*ct*_ = (*x*_*ct*1_ … *x*_*ctp*_) is a vector of *p* covariates for county *c* during year *t*, with *c = 1, …, C* for the *C* counties in the US and *t = 1,…,N*, for *N* years in the study period; ***β*** = (*β*_1_ … *β*_*p*_) is the vector of covariate regression parameters whose Gaussian prior is defined by mean µ_*β*_ and diagonal variance matrix ***Σ***_*β*_. *ψ*_*ct*_ a latent component encompassing one or more sets of spatiotemporally autocorrelated random effects. The CAR-ANOVA(47) model decomposes spatiotemporal variation, *ψ*_*ct*_, into an overall spatial effect across the study period (***ϕ***), an overall temporal trend over the study area (***δ***), and a set of independent space-time interactions (***γ***); ***ϕ*** = (*ϕ*_1_ … *ϕ*_*C*_) and ***δ*** = (*δ*_1_ … *δ*_*N*_) are modeled by the CAR prior proposed by Leroux and others (34).

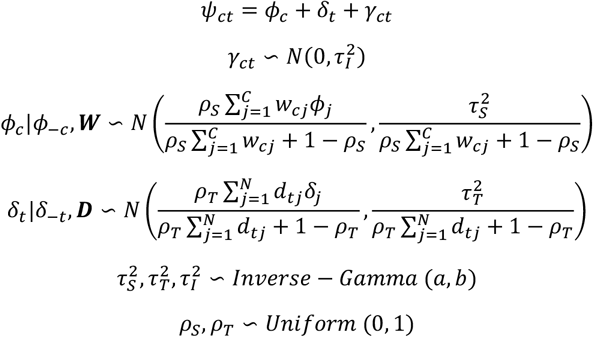

Here, **W** is the *C* x *C* spatial adjacency matrix, with *wcd* = 1 if counties *c* and *d* are adjacent to each other and 0 otherwise (counties are adjacent if they share at least one boundary point in the shape file; a county is not adjacent to itself). Analogously, ***D*** is the *N x N* temporal adjacency matrix, with *dtj* = 1 if |*t - j*| = 1 (i.e. consecutive years) and 0 otherwise. The priors for the spatial 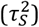, temporal 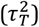 and space-time interaction 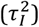 random effects variances are specified by an Inverse-Gamma distribution with *a=1* and *b=0*.*01*; spatial (*ρ*_*S*_) and temporal (*ρ*_*T*_) dependence parameters have uniform priors in the unit interval (1 indicates strong dependence; 0 independence).

The models were fit in a Bayesian setting using Markov chain Monte Carlo simulations. Parameters whose full conditional distributions have a closed form distribution are Gibbs sampled and the rest are updated using the Metropolis adjusted Langevin algorithm (48). For each model, three Markov chains each 110,000 in length with a burn-in of 10,000 were generated and a thinning factor of 1,000 was applied to remove correlation among the samples (49-51). Convergence was verified with the Geweke diagnostic statistic (52). Implementations of the methods are per CARBayesST package (53, 54) in R (55).

### Statistical Analysis and Evaluation

To ascertain that methods that explicitly accommodate spatiotemporal autocorrelation are necessary, we initially built Poisson log-linear models and verified the presence of autocorrelation in their residuals using the Moran’s I statistic (56). Further, in the interest of model parsimony, to identify variables with marginal contribution to model quality, we built log-linear models with all possible combinations of predictors considered (2^7^ – 1) and compared their goodness-of-fit (Akaike Information Criterion (AIC)) against that of a model built using all available predictors. A model with a subset of 5 predictors (excluding weekly wage, unemployment rate, and depression prevalence variables) was found to have an AIC very close to the full model.

Subsequently, we built CAR-ANOVA models with: i) the *full* set of predictors; ii) the *select* subset of predictors; and iii) no covariates i.e. a *null* model, to measure the predictive skill from accounting for autocorrelation alone. In addition, a *reference* model, with the expected deaths in a county estimated from differential risk by age, race, and sex of the county’s population was also used (see Appendix Text 2). This model did not capture spatial patterns or temporal trends in suicide mortality and provides a benchmark estimate to assess improvements from CAR models. The *select* model is our primary model in this study and all reported findings, unless otherwise stated, are based on its estimates.

To estimate yearly risk over the study period, we computed average risk across all counties for each MCMC sample, and report median and 95% interval ranges over all samples. Estimates of yearly county-specific risk are median over all samples’ estimates for the specific year and county.

Outcome and predictor data overlap for the years 2005-2016. We defined this as our study period, and all models were trained on data for these years. As mortality outcome data and most of the predictors are available for three additional years (2017-2019), we use these surplus years’ data to calculate the out-of-sample (OOS) predictive model skill, by assuming unavailable predictors remain unchanged since their last known values. For temporal OOS validation, risk for a given year is predicted with models fit using data up to, but not including, the given year. For spatial OOS validation we used 10-fold cross validation: partitioned the counties into ten roughly equal folds, trained the models 10 times with one partition of the counties withheld in each iteration, and used the fit models to predict risk in holdout counties. We compared the model errors over the fit period (2005-2016) and under the two OOS settings, temporal (2017-2019) and spatial.

Errors were calculated using the symmetric proportional error (SPE), 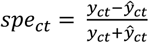, where *yct* and ŷ denote observed and predicted deaths, respectively. SPE has a well-defined range and indicates the direction of the error. A division-by-zero issue was avoided by imposing a small lower bound on ŷ. An aggregate measure of SPE, the Mean SPE (MSPE), was used to compare model accuracy on both in-sample and OOS predictions. The mean in-sample error for a model was calculated as 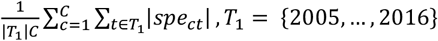, the mean temporal OOS error as 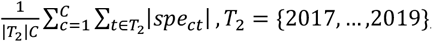, and spatial OOS error as 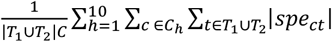, where *C* _h_ denotes counties held out in fold *h*. Wilcoxon signed rank test was used to assess statistical significance in the difference of errors for each pair of model forms (57).

## Results

Figure 1 shows the median and 95% credible interval of the posterior effect estimates for each standard deviation change in predictor value (see Appendix Table 1 for values). Suicide mortality risk is 2.8% higher for each 11% increase in state’s firearm ownership rate and by 1% for each 0.7% increase in prevalence of major depressive disorder in the state. Conversely, risk was lower by 4.3% for each $12,000 increase in annual median household income in the county, and by 4.3% for each 5.8 increase in population per square mile in the county (see Appendix Table 2 for effect estimates per unit change in predictor value). A not statistically significant decrease of 0.3% for each $162 of weekly wage was also estimated.

**Figure 1.**
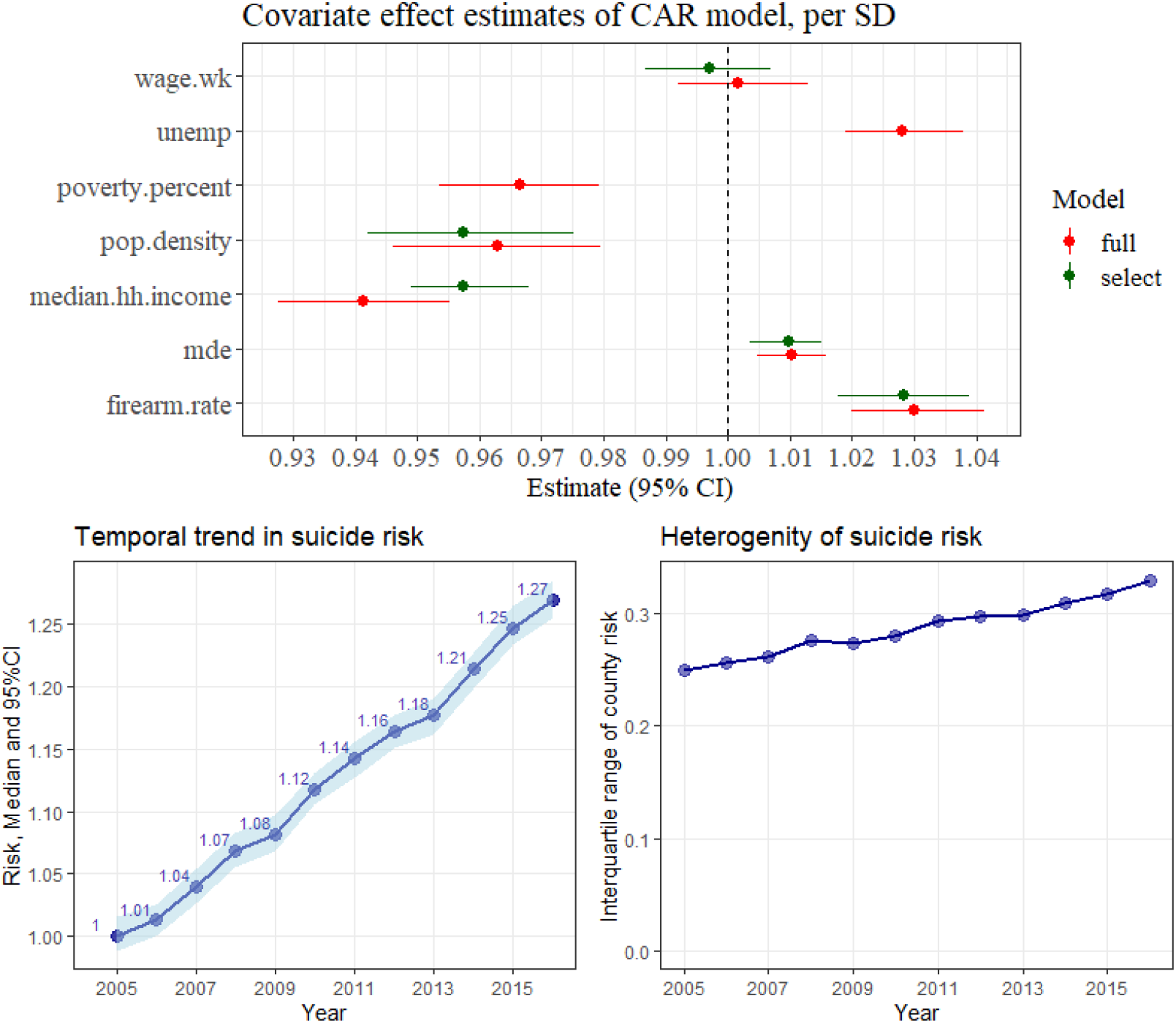
(*A: top*) Posterior median and 95% credible intervals of the effect estimates from CAR-ANOVA models, for one standard deviation change in variable. The variable descriptions are listed in Table 1.; (*B: bottom, left*)Median and 95% CI for national suicide risk; (*C: bottom, right*) Interquartile (IQR) range of county-level risk as estimated by the *select* model. Higher IQR indicates greater heterogeneity and an increase in IQR with time indicates a widening gap between low and high-risk counties.

The protective effect of poverty seen in the *full* model (lower by 3.4% for each 6.4% increase in poverty prevalence in the county) needs more careful examination and interpretation, ideally in conjunction with measures of rurality, societal fragmentation and poverty persistence (58, 59) and interactions with other predictors in the model. The findings are not anomalous however, as previous studies have reported mixed associations between poverty prevalence and suicide rates at the population-level, with results varying by study design including geographical resolution and population strata analyzed (22). At the individual-level, however, the negative impact of poverty on suicide rates is more consistent across studies (60, 61).

Overall, the effect estimates of the variables in the *select* model remained largely unchanged when additional covariates were introduced i.e. in the *full* model (Figure 1A). Both the *select* and *full* models also detected strong spatial dependence (*ρ*_*S*_ = 0.97) and temporal dependence (*ρ*_*T*_ = 0.91) (Appendix Table 2).

### Heterogeneity in suicide risk

The model’s estimate of annual average suicide risk nationally showed a clear increasing trend from 1 in 2005 to 1.27 in 2016, with smaller increases in 2009, immediately after the onset of the 2008 recession, and in 2013 (Figure 1B). This trend is consistent with increases in mortality rates reported by multiple studies for the US overall and in almost all demographic groups (1).

Results also showed an increase in the heterogeneity in county-specific suicide risk during the study period (Figure 1C), indicative of a widening gap between low-risk counties and high-risk counties. Neither trend estimate was found to be sensitive to the set of covariates included in the model (Appendix Figure 2). Although beyond the scope of our current analysis, these county-specific risk estimates can help categorize counties — for example, counties with relatively stable risk especially those that remained in the highest or lowest deciles, or counties that experienced the largest year-to-year changes — and hence help identify areas in greater need of preventive resources, or conversely identify areas where interventions appear to be effective.

### Comparison of model errors

Figure 2 shows that the *select* model’s predictive ability was comparable to that of the *full* model, with in-sample errors of *select* smaller than *full*, spatial OOS errors larger and temporal OOS not significantly different (*p* = 0.46). This finding implies that two of the covariates did not contribute to model quality in the presence of the other predictors, possibly due to collinearities, yielding a more parsimonious model dependent on fewer data sources.

**Figure 2.**
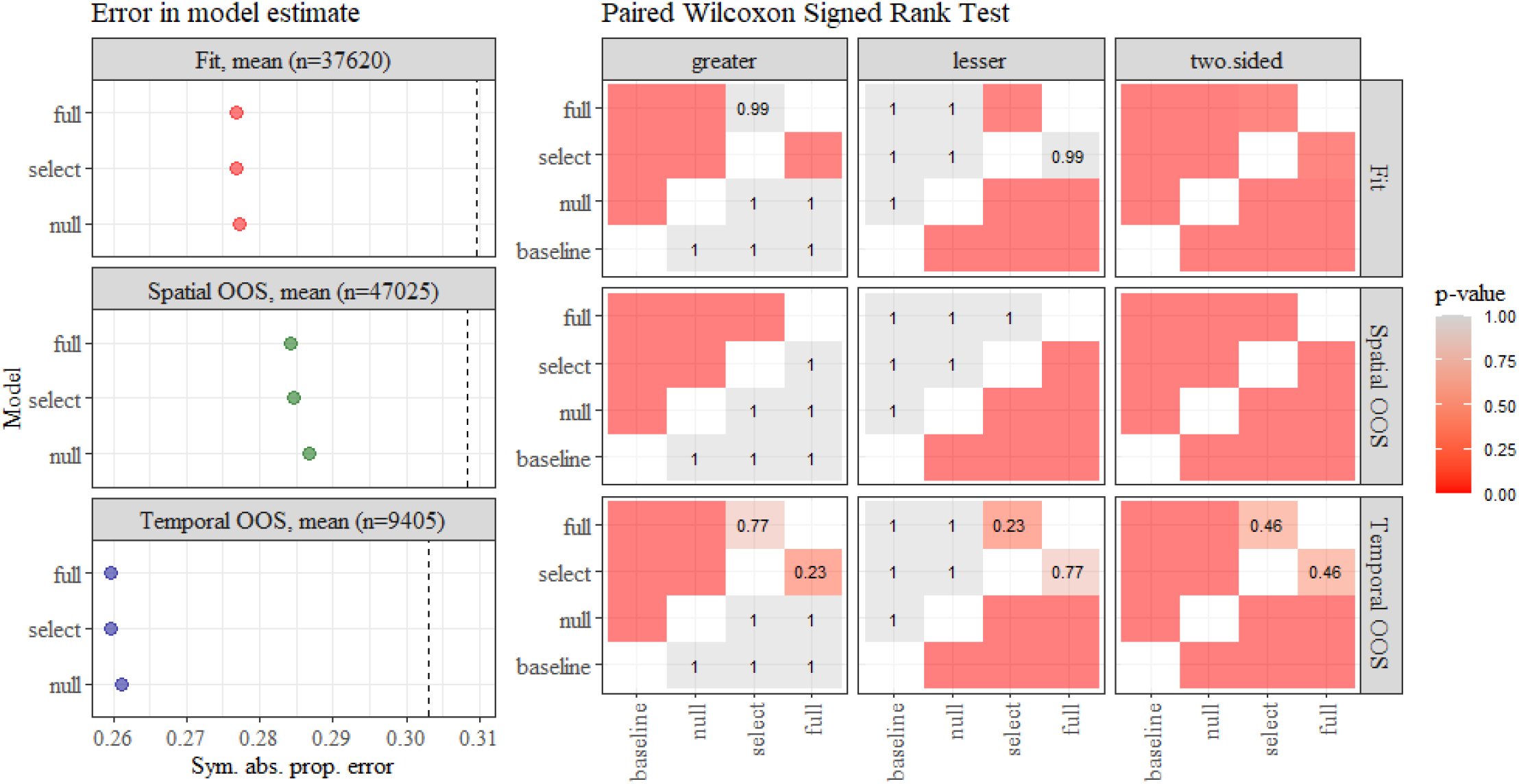
(*left*) Mean symmetric proportional error for in-sample, temporal out-of-sample and spatial out-of-sample estimates. The vertical dashed line denotes error from the *reference* model; (*right*) Wilcoxon signed rank test for each pair of models. Significance (*p* < .05) in the ‘two.sided’ column indicates that the difference in errors of Model *X* and Model *Y* is not symmetric around 0. *p* < .05 in the ‘lesser’ panel column indicates errors in Model *X (x-axis)* are lower than Model *Y*. Actual *p* -value are shown as text when errors are not significantly different. To interpret this plot, for a pair of models, verify that the difference is significant under the ‘*two-sided’* test, and if true, check for significance under the ‘*lesser*’ or ‘*greater’* panel columns.

Both the *select* and *full* models had lower error than the *null* model in all three settings (fit, temporal and spatial OOS) and the differences were statistically significant (*p* < .05), indicating that the inclusion of covariates improves the model over what is gained by accounting for spatiotemporal autocorrelation in suicide mortality alone. Furthermore, all three CAR-ANOVA models had lower errors than the *reference* model, indicating an improved predictive ability from including spatiotemporal associations. Errors from the *reference* model were 7%-14% larger than errors from the *null* model (Appendix Figure 3), suggesting that the CAR-ANOVA models could be of value even in settings where SES predictors of suicide mortality are not available.

Geweke and Gelman-Rubin diagnostic tests (Appendix Table 3) and visual inspection of trace plots (Appendix Figure 4) indicated model convergence. Scatter plot of predicted suicide deaths against observed deaths in the temporal OOS period, showed possible overprediction at lower counts (Appendix Figure 5) but otherwise reasonable estimates.

### Autocorrelation in residuals

As a primary motivation for the use of the CAR models was to account for spatiotemporal autocorrelation, to test whether the CAR model form was adequate to capture autocorrelation, we looked for autocorrelation in models’ residuals. A visual inspection of the spatial distribution of in-sample (Figure 3) and temporal OOS residuals (Figure 4, bottom row) showed no clear spatial structure. Formal Moran’s I test found no statistically significant spatial autocorrelation in spatial OOS setting for any year; however, for a majority of the years in both in-sample and temporal OOS settings significant autocorrelation was detected (*p* < .05). Reassuringly however, the magnitude of the autocorrelation with the CAR-ANOVA models, was considerably lower than the *reference* (Appendix Figure 6).

**Figure 3.**
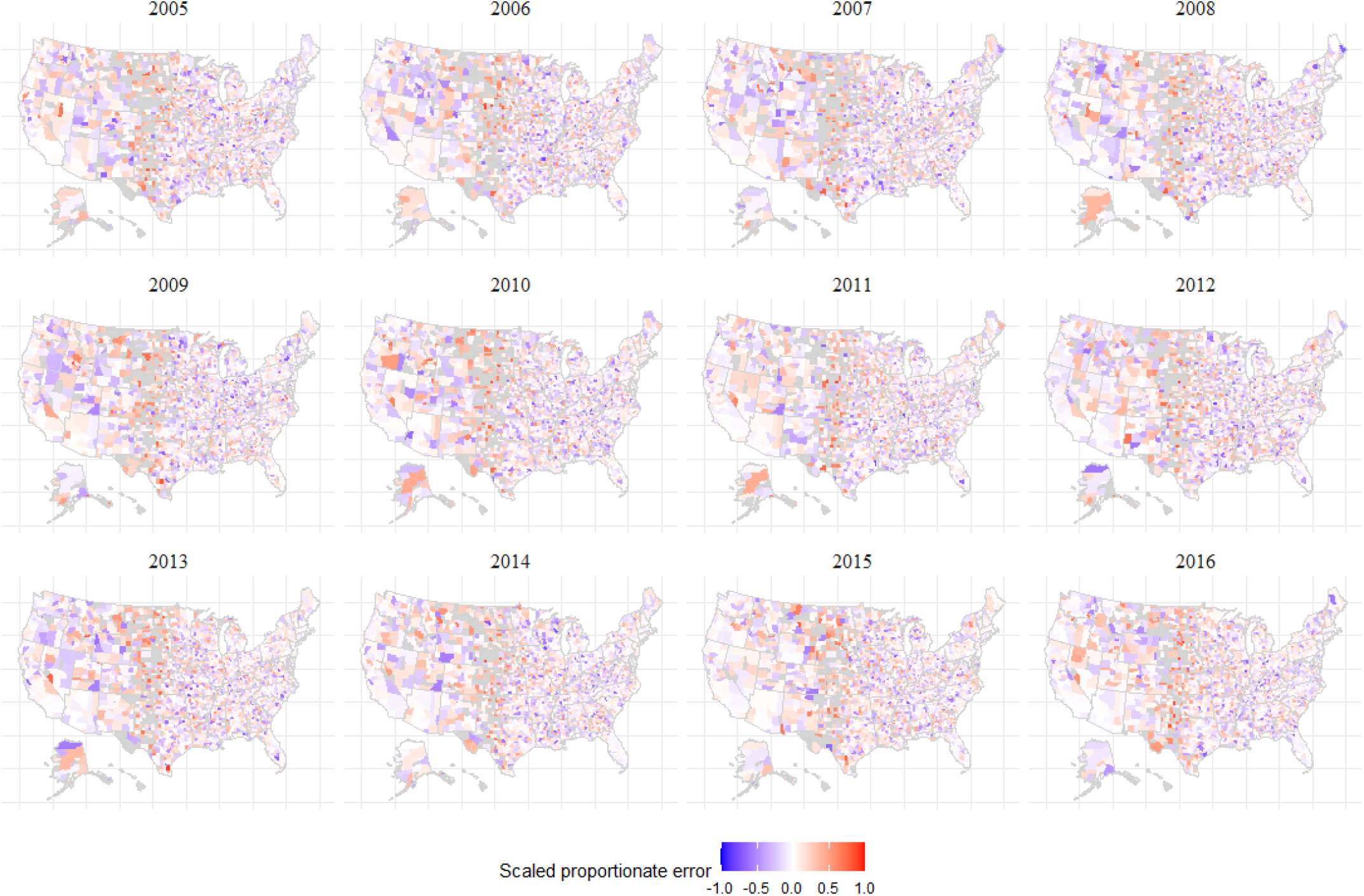
Symmetric proportional in-sample errors for *select* model. Negative error indicates model estimate exceeds observed. Counties with no observed deaths are shown in grey.

**Figure 4.**
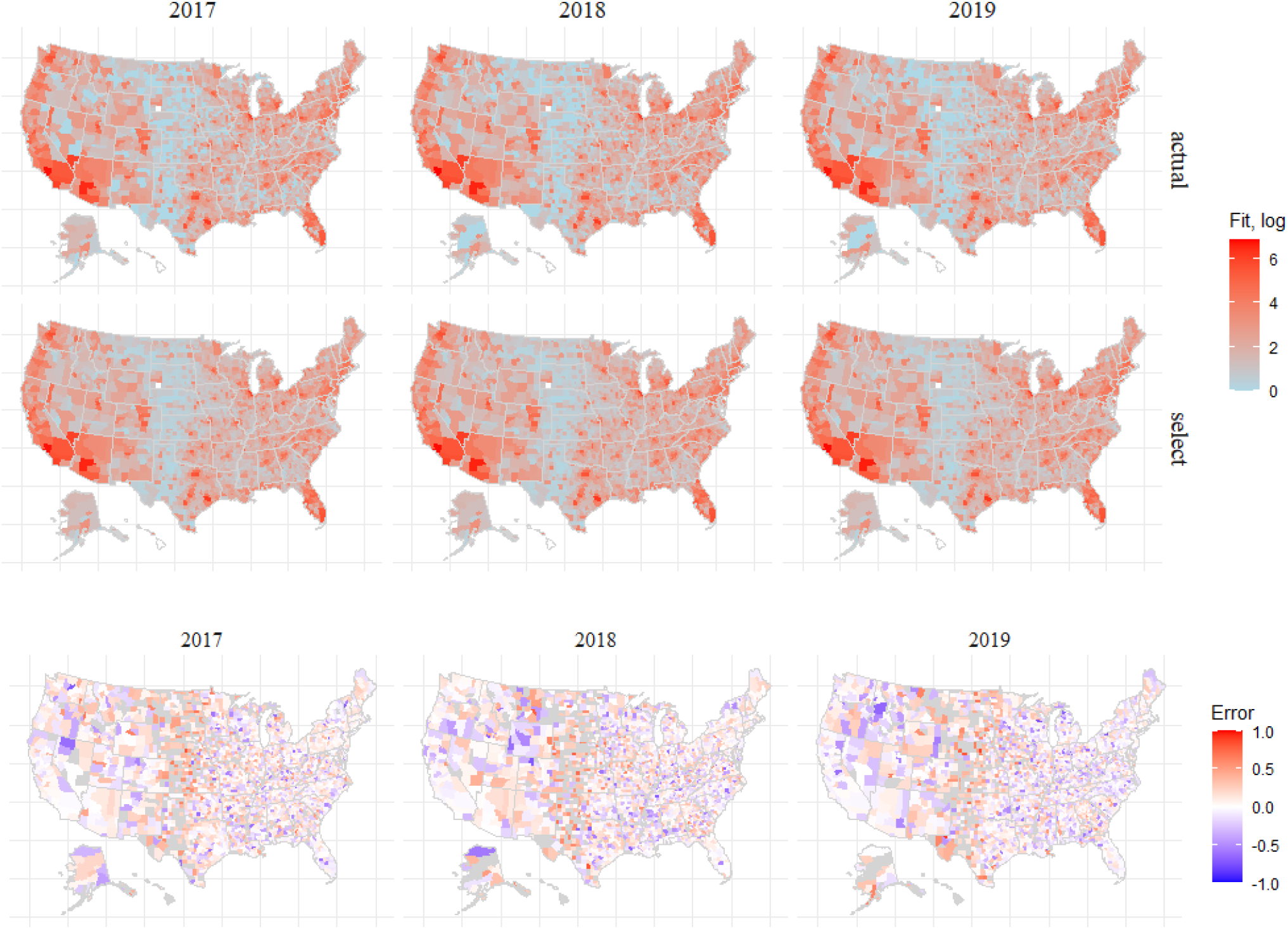
Observed suicide deaths (top row), out-of-sample estimates (middle row) and symmetric proportional errors for *select* model. Negative error indicates model estimate exceeds observed. Counties with no observed deaths are show in grey.

This reduction but not elimination of spatial autocorrelation could be indicative of the insufficiency of the specific CAR model form used and/or of the covariates considered. In addition, two of the predictors — prevalence of major depressive episode and firearm ownership — were only available at state-wise resolution, a shortcoming addressed by assuming all counties in a state to have identical values, which may have contributed to an increase in residual spatial autocorrelation.

## Discussion

Prediction models of suicide typically only consider clinical characteristics in high-risk clinical population and general population settings, yet suicide risk is also spatially and temporally determined. Our results demonstrate that predictions of suicide death are improved when aspects of the social environment are used to model risk, and thus aid suicide prevention efforts. Absence of evidence of over-fitting in out-of-sample validation lends confidence to future risk estimates with these models.

Given that the primary objective of this study was an evaluation of the feasibility of a predictive model, the covariates considered are not exhaustive. A more comprehensive review of domain literature can help identify a more robust set of predictors not collinear with the predictors considered (62). Additional stratifications of the current predictors or other SES indicators, such as measures of social cohesion, access to healthcare, unemployment rates in specific sectors of the economy, as well as prevalence of depression stratified by age, prescription rates for antidepressant and pain management medications and housing quality (crowded living conditions, access to green spaces, ambient vehicular noise) could also be tested. When used at sub-national scales by a state or local public health agency, additional predictors of local relevance that do not have national coverage may also be viable.

The predictions from the models presented here are at annual resolution and hence not responsive to real-time changes in risk. While the model structure does not preclude generation of weekly or monthly risk estimates, a barrier to such an operational deployment is the paucity of reliable, timely measures of suicidal activity (thoughts or attempts). Identification of sources for near real-time situational data, and development of nowcast models to translate data feeds into measures of suicidal ideation in the community, could significantly aid the translation of the models discussed here into a surveillance setting.

One potentially good source of information on ideation is the volume of calls to crisis hotline centers. With the forthcoming launch of a national suicide prevention number (9-8-8) in mid-2022, a unified system with wide coverage is at hand. Similarly, with the deployment of the National Emergency Medical Services Information System (NEMSIS), timely information on EMS requests is also available (63, 64). Aggregate event data at county resolution from these and/or similar sources, if made publicly available every week, could support the development of nowcast systems. Such models are in use in numerical weather prediction (65, 66), macroeconomic analyses (67, 68) and influenza surveillance (69-71), among other domains.

Indicators of suicidal crisis or ideation may also be inferred from posts to thematically-related social media sites (72-76), queries on search engines (77) or access logs to suicide prevention forums and related websites. Sources of ideation among teens and young adults, among whom suicide is in the top 3 common causes of death, could be particularly valuable (78). Coupled with such measures of situational awareness, the CAR models might flag anomalous changes in population-level suicide risk, and thus inform decisions on resource allocation and focused interventions during ongoing outbreaks. Reducing suicide deaths, alongside unintentional drug overdose deaths, may prove critical to public health efforts aimed at reversing declines in life expectancy in the US.

Limitations of the study include *ad hoc* independent variable selection, the assumption of geographical homogeneity for predictors lacking county-level data, a potentially simplistic spatial adjacency matrix that did not reflect population mobility and mixing, inflated error estimates in zero-death counties (see Appendix Text 3), and potential underestimates of suicide deaths and risk in certain racial/ethnic groups due to inconsistent suicide certification practices (79).

## Data Availability

Covariate data were gathered from public datasets and references to the primary sources have been provided in the manuscript. Mortality data was acquired under a restricted use agreement that does not allow sharing data publicly.

## Acknowledgements

This work is funded by a grant from the National Institute of Mental Health (R01-MH121410) to Keyes and Shaman. The funder had no role in study design; collection, analysis, and interpretation of data; writing the report; and the decision to submit for publication.

## Data/code sharing statement

All statistical code used to run the models and perform analysis can be shared. Covariates were gathered from public datasets and references to the primary sources have been provided. Mortality data was acquired under a restricted use agreement that does not allow sharing data publicly.

## Author Contributions

Designed research: Keyes, Shaman, Kandula

Performed research: all authors

Analyzed data and developed tools: Gimbrone, Rutherford, Kandula

Wrote the paper: (initial draft) Martinez-Alés, Kandula; (revision) all authors

