## Supporting text and figures for "Modeling suicide mortality in US counties using population socioeconomic indicators"

**This PDF includes:**

Appendix text, 3 sections
Appendix Tables 1 - 3
Appendix Figures 1 - 7

***Appendix Text 1: Detailed description of independent variables***

*Proportion of population living in poverty; median household Income*

US Census Bureau's Small Area Income and Poverty Estimates (SAIPE) program (37) provides annual estimates of measures of income and poverty at small geographical resolutions (county and school district), which are either unavailable or only available infrequently. For example, the 1-year American Community Survey (ACS) provides data on persons living in poverty only for counties with a population greater than 65000. SAIPE uses a regression model to estimate poverty rates in all counties with the number of persons in poverty as the dependent variable (in counties where ACS survey estimates are available) and multiple predictor variables from the Supplemental Nutrition Assistance Program (SNAP), federal income tax returns and US Decennial census. Estimates of median household income are similarly obtained from a separate regression model with additional predictor variables provided by the Bureau of Economic Analysis.

*Prevalence of major depressive episodes*

The National Surveys on Drug Use and Health (NSDUH) dataset contains state-level small area estimates on key substance use and mental health outcomes (38). The depression prevalence indicator is the estimated proportion of population with at least one major depressive episode during the previous year. As county-level estimates of prevalence are not available from this data source and we are unaware of other reliable sources, the annual prevalence is assumed to be the same in all counties of a state.

*State prevalence of firearm-owning households*

RAND's Household Firearm Ownership Database (39) provides annual estimates of the proportion of adults who live in a household with firearms for each state in the US between 1980 and 2016. These estimates are based on direct measures of ownership from individual-level survey data and indirect proxy measures of ownership (for example, per capita hunting licenses, background checks, and subscriptions to *Guns & Ammo* magazine).

As the surveys were designed to be nationally representative but not for each state, multi-level regression with post-stratification was used to calculate subnational estimates (A1). These corrected direct measures are then combined with indirect measures using a structural equation model which attempts to represent both direct and indirect measures as dependent in part on household ownership rates and in part on observed and unobserved confounders.

Additionally, as one of the indirect measures used by RAND (proportion of suicides that involved firearms) is collinear with the outcome of interest in this study, we re-estimated the firearm ownership rates with this measure excluded. Household firearm ownership rates were assumed to be homogenous across all counties in the state, a necessary simplification given annual county-level estimates of firearm ownership are unavailable.

*Average weekly wage*

The Bureau of Labor Statistics through the Quarterly Census of Employment and Wages program (40) provides timely and finely-resolved estimates of wages from several industries covering over 95% of US jobs. These estimates are based on employer reports to the unemployment insurance contribution system and two annual surveys. Data are aggregated to industry sectors and geographic levels (metro, county, state, and national) and are available at annual frequency, with more frequent releases available at higher aggregations. Here, we use county-wise estimates of annual average weekly wage across all industries.

*Unemployment rate*

The Bureau of Labor Statistics through the Labor and Unemployment Statistics (41) program provides estimates of unemployment by combining data from the Current Population Survey, the Current Employment Statistics survey, and state unemployment insurance systems. County-level rates incorporate methodological corrections to include agricultural workers, self-employed, unpaid family workers and private household workers who are not otherwise represented in administrative/survey datasets. These estimates are considered reliable and form the basis for budgetary allocations by federal, state and local governments.

*Population Density*

Annual population density in each county was estimated using the intercensal and postcensal population estimates described above and the county land area per the 2010 US census (42). This calculation is not sensitive to changes in county boundaries during the study period. A log transformation was applied as the distribution was found to be non-normal.

Appendix Figure 1 shows pairwise Spearman correlation for each pair of covariates.

***Appendix Text 2: Reference model***

Let $y_{ct}$ denote observed count of suicide deaths in county $c$ during year $t$, and $p_{ct}$the corresponding population estimate. We assume the population can be split into $k$ mutually exclusive and exhaustive strata, each with a different risk of suicide death; let $y_{ctk}$, $p_{ctk}$ be the respective suicide deaths and population in strata $k$. The suicide risk for strata $k$ nationally over the study period (*t=1…n*) is calculated as:

$$\eta_k = \frac{1}{n}\sum_{t=1}^{n}\left(\frac{\sum_c y_{ctk}}{\sum_c p_{ctk}}\right)$$

The expected number of deaths in county $c$ during year $t$ is calculated as:

$$\mu_{ct} = \sum_k p_{ctk} * \eta_k$$

Drawing on prior studies on heterogeneity of suicide risk by age, race and gender, we use $k$ = 72 strata of 9 age groups ([5, 15), [15, 25), …, 85+), 4 racial groups (White, Black, American Indian/Alaskan Native, and Asian/Pacific Islander) and two gender groups. The race and gender categories are identical to those provided by the bridged-race population datasets.

***Appendix Text 3: Isolated counties and counties with no recorded deaths***

Of the 3142 counties in the US, 5 counties (Honolulu, HI; Kauai, HI; Hawaii, HI; Nantucket, MA; and San Juan WA) have no known neighbors in the shape file. As the CAR model specification requires each location to have at least one neighbor, these counties were excluded from the analysis. Additionally two counties (Wade Hampton, AK and Shannon, SD) that had no outcome data were also excluded.

There were also a sizeable number of counties with no recorded deaths during a year. Over the 12 year study period, a total of 4979 county-year instances (13.2% of total) had zero suicide death counts, with the yearly percentages decreasing from 15.2% in 2005 to 10.6% in 2016. When the observed outcome is 0, the symmetric proportion error used as the error measure in this study, takes the maximum possible penalty of 1, considerably inflating the aggregate mean errors. As we are unaware of a consensus on how to handle these zero-count cases, we retained these counties while calculating the errors in the main text. Appendix Figure 7 shows the errors with these county-year instances excluded. Dropping these county-year pairs does not change the improvement in errors of all CAR-ANOVA models relative to the *reference* model.

| Variable | Select | Full |
|---|---|---|
| Firearm ownership | 1.028 (1.02, 1.04) | 1.03 (1.02, 1.04) |
| Major Depression, % | 1.01 (1, 1.02) | 1.01 (1, 1.02) |
| Median HH income | 0.957 (0.95, 0.97) | 0.941 (0.93, 0.96) |
| Population density | 0.957 (0.94, 0.98) | 0.963 (0.95, 0.98) |
| Poverty, % | NA | 0.966 (0.95, 0.98) |
| Unemployment, % | NA | 1.028 (1.02, 1.04) |
| Weekly wage | 0.997 (0.99, 1.01) | 1.002 (0.99, 1.01) |

**Appendix Table 1**. Effect estimates for suicide mortality risk for one standard deviation change in predictor.

| Variable | Mean Estimate | 95% CI |
|---|---|---|
| Firearm ownership | 0.246 | 0.16, 0.34 |
| Major Depression, % | 1.455 | 0.56, 2.19 |
| Median HH income | -3.681 | -4.42, -2.78 |
| Population density | -0.024 | -0.03, -0.02 |
| Weekly wage | -0.019 | -0.08, 0.04 |
| rho.S | 0.973 | 0.94, 0.99 |
| rho.T | 0.911 | 0.61, 0.99 |
| tau2.I | 0.003 | 0.002, 0.004 |
| tau2.S | 0.099 | 0.091, 0.108 |
| tau2.T | 0.003 | 0.001, 0.007 |
| WAIC | 161235 | |

a. *Select* model

| Variable | Mean Estimate | 95% CI |
|---|---|---|
| Firearm ownership | 0.262 | 0.18, 0.36 |
| Major Depression, % | 1.500 | 0.73, 2.31 |
| Median HH income | -5.149 | -6.37, -3.96 |
| Population density | -0.022 | -0.03, -0.01 |
| Poverty, % | -0.533 | -0.74, -0.34 |
| Unemployment, % | 0.954 | 0.66, 1.27 |
| Weekly wage | 0.011 | -0.05, 0.08 |
| rho.S | 0.975 | 0.94, 0.99 |
| rho.T | 0.914 | 0.61, 0.99 |
| tau2.I | 0.003 | 0.002, 0.004 |
| tau2.S | 0.096 | 0.088, 0.104 |
| tau2.T | 0.003 | 0.002, 0.008 |
| WAIC | 161250 | |

b. *Full* model

| Variable | Mean Estimate | 95% CI |
|---|---|---|
| rho.S | 0.982 | 0.958, 0.995 |
| rho.T | 0.893 | 0.572, 0.987 |
| tau2.I | 0.003 | 0.002, 0.004 |
| tau2.S | 0.120 | 0.111, 0.129 |
| tau2.T | 0.002 | 0.001, 0.006 |
| WAIC | 161267 | |

c. *null* model

**Appendix Table 2**. Effect estimates in suicide mortality risk for one *unit change* in predictor, and spatial and temporal dependence parameters. WAIC: Watanabe-Akaike Information Criterion

**Chain 1**: WAIC= 161229;DIC = 161100; p.d. = 2613; LMPL = -80634

| | Median | 2.5% | 97.5% | n.effective | Geweke.diag |
|---|---|---|---|---|---|
| (Intercept) | 0.1505 | 0.0683 | 0.2427 | 100 | -0.2 |
| Weekly wage | -0.0189 | -0.0818 | 0.045 | 100 | -0.6 |
| Median HH income | -3.5928 | -4.4867 | -2.6009 | 100 | 0.9 |
| Major depression, % | 1.4582 | 0.5275 | 2.1948 | 100 | 0.1 |
| Firearm ownership | 0.2369 | 0.1578 | 0.3238 | 149 | 0.9 |
| Population density | -0.0252 | -0.0323 | -0.0154 | 73.6 | -0.9 |
| tau2.S | 0.0991 | 0.0909 | 0.1081 | 100 | 0.5 |
| tau2.T | 0.0031 | 0.0012 | 0.0069 | 100 | -0.7 |
| tau2.I | 0.0029 | 0.0022 | 0.0037 | 23 | 0.7 |
| rho.S | 0.9737 | 0.9466 | 0.9912 | 100 | -0.3 |
| rho.T | 0.8983 | 0.5141 | 0.9902 | 100 | 0.6 |

**Chain 2**: WAIC= 161239; DIC = 161107; p.d. = 2619; LMPL = -80641

| | Median | 2.5% | 97.5% | n.effective | Geweke.diag |
|---|---|---|---|---|---|
| (Intercept) | 0.1447 | 0.0804 | 0.2158 | 100 | -1.9 |
| Weekly wage | -0.0166 | -0.084 | 0.0357 | 100 | -0.8 |
| Median HH income | -3.7457 | -4.4904 | -2.7739 | 100 | 1.5 |
| Major depression, % | 1.5112 | 0.6544 | 2.263 | 100 | -0.7 |
| Firearm ownership | 0.2527 | 0.1689 | 0.3581 | 100 | 1.3 |
| Population density | -0.0244 | -0.0331 | -0.0162 | 100 | 2.6 |
| tau2.S | 0.0989 | 0.0913 | 0.108 | 100 | 2.1 |
| tau2.T | 0.0025 | 0.0013 | 0.0074 | 100 | -0.1 |
| tau2.I | 0.003 | 0.0024 | 0.0037 | 49.2 | 1.1 |
| rho.S | 0.974 | 0.9379 | 0.9915 | 70.2 | 1.7 |
| rho.T | 0.9192 | 0.74 | 0.994 | 100 | 1.3 |

**Chain 3**: WAIC= 161239;DIC = 161115; p.d. = 2633; LMPL = -80645

| | Median | 2.50% | 97.50% | n.effective | Geweke.diag |
|---|---|---|---|---|---|
| (Intercept) | 0.1534 | 0.0652 | 0.246 | 584 | 0.3 |
| Weekly wage | -0.0203 | -0.0793 | 0.0409 | 100 | -0.4 |
| Median HH income | -3.7046 | -4.2836 | -2.9766 | 100 | 0.8 |
| Major depression, % | 1.3949 | 0.4921 | 2.1189 | 100 | -1.1 |
| Firearm ownership | 0.248 | 0.1489 | 0.3352 | 100 | 0.5 |
| Population density | -0.0239 | -0.0358 | -0.0138 | 100 | -0.3 |
| tau2.S | 0.0986 | 0.0903 | 0.1081 | 100 | 0.9 |
| tau2.T | 0.0029 | 0.0014 | 0.0065 | 100 | 0.4 |
| tau2.I | 0.003 | 0.0023 | 0.0037 | 50.3 | 0.6 |
| rho.S | 0.9712 | 0.9425 | 0.9902 | 100 | -2.5 |
| rho.T | 0.9146 | 0.588 | 0.9899 | 100 | -0.1 |

*Gelman-Rubin Diagnostic*: 1.02

**Appendix Table 3**: Output from the *select* model over the fit period from three chains; showing mean and 95% credible interval; effective number of independent samples and diagnostics. Geweke diagnostic between [-2, 2] indicates model convergence and Gelman-Rubin statistic under 1.1 indicates that longer chains are not necessary. *WAIC*: Watanabe-Akaike Information Criterion *DIC*: deviance information criteria; *p.d.*: effective number of parameters; *LMPL*: Log Marginal Predictive Likelihood.

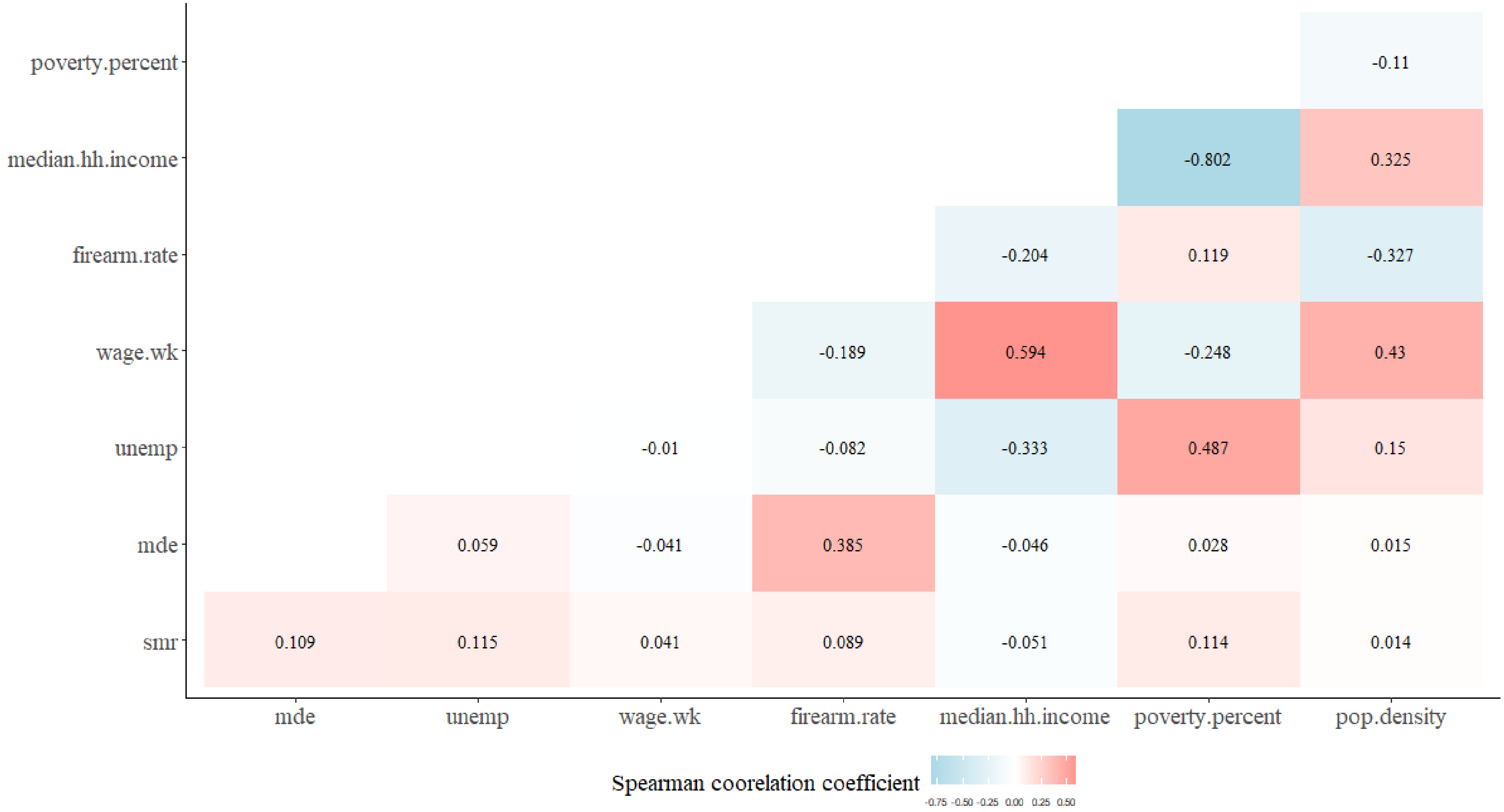


**Appendix Figure 1.** Spearman correlation coefficient for variables used. *smr* = standardized mortality rate; *mde*: major depressive episode; *unemp*: unemployment rate; *wage.wk*: weekly wage; *median.hh.income*: median household income

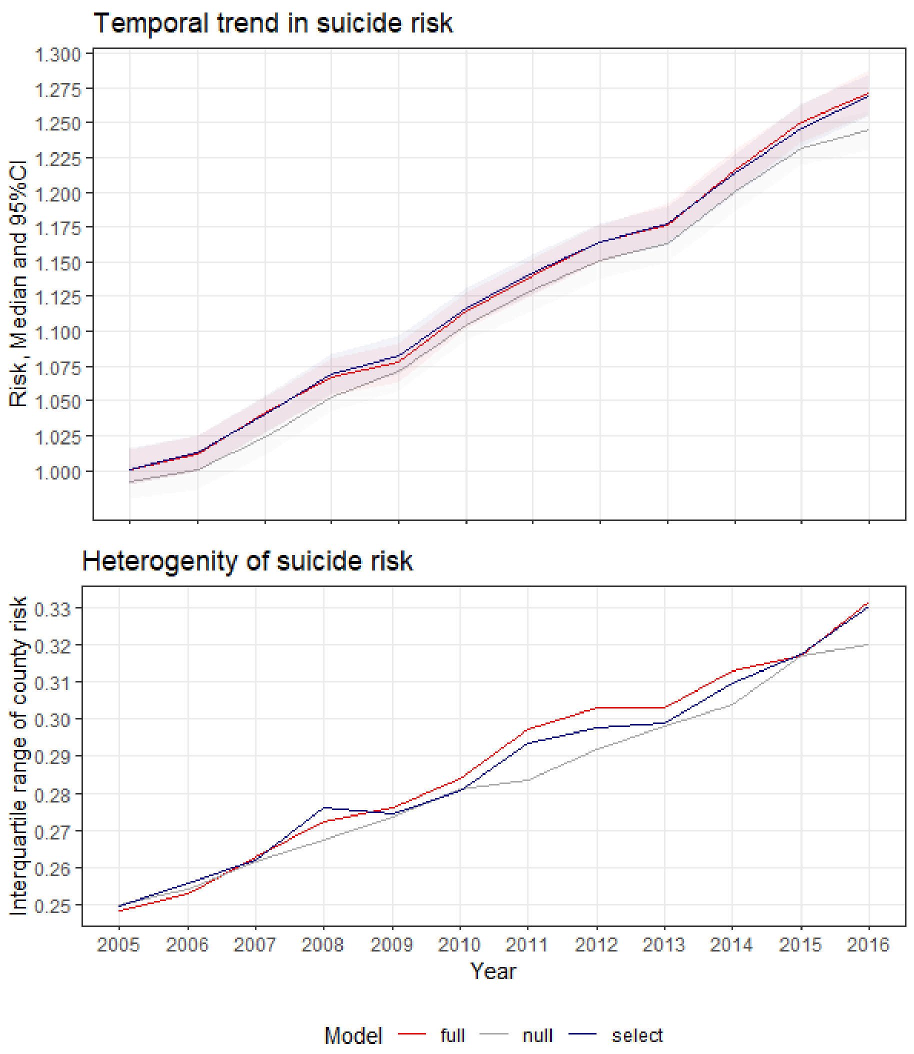


**Appendix Figure 2.** a) Median and 95% CI for national suicide risk for the three CAR models (risks from *select* and *full* models are similar and not distinguishable). B) Interquartile (IQR) range of county-level risk for the same models.

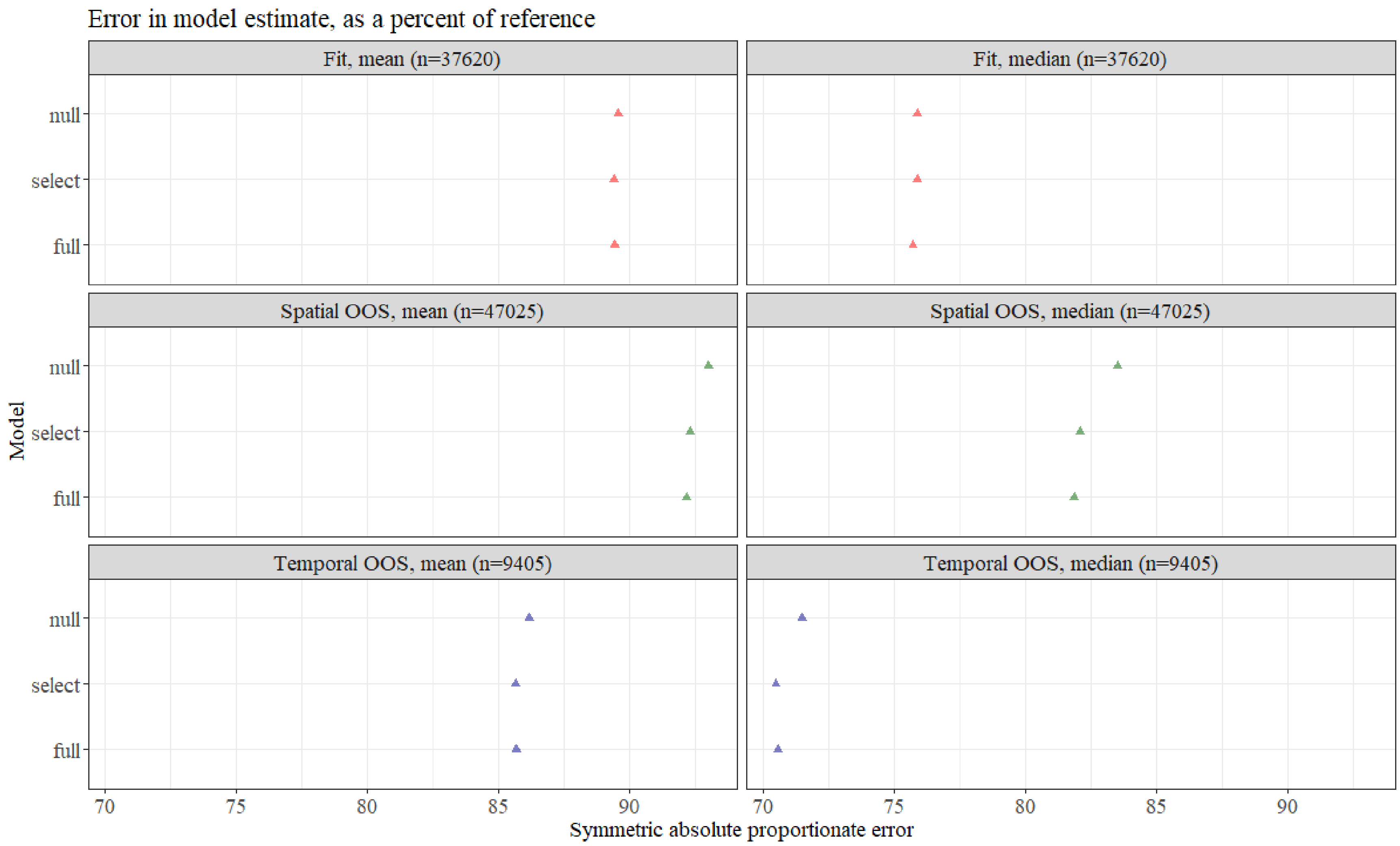


**Appendix Figure 3.** Mean and Median symmetric proportional error for in-sample, temporal out-of-sample and spatial out-of-sample estimates for the three CAR models, as a percent of error in *baseline*. Baseline estimates are expected deaths from population profile, $\mu_{ct}$.

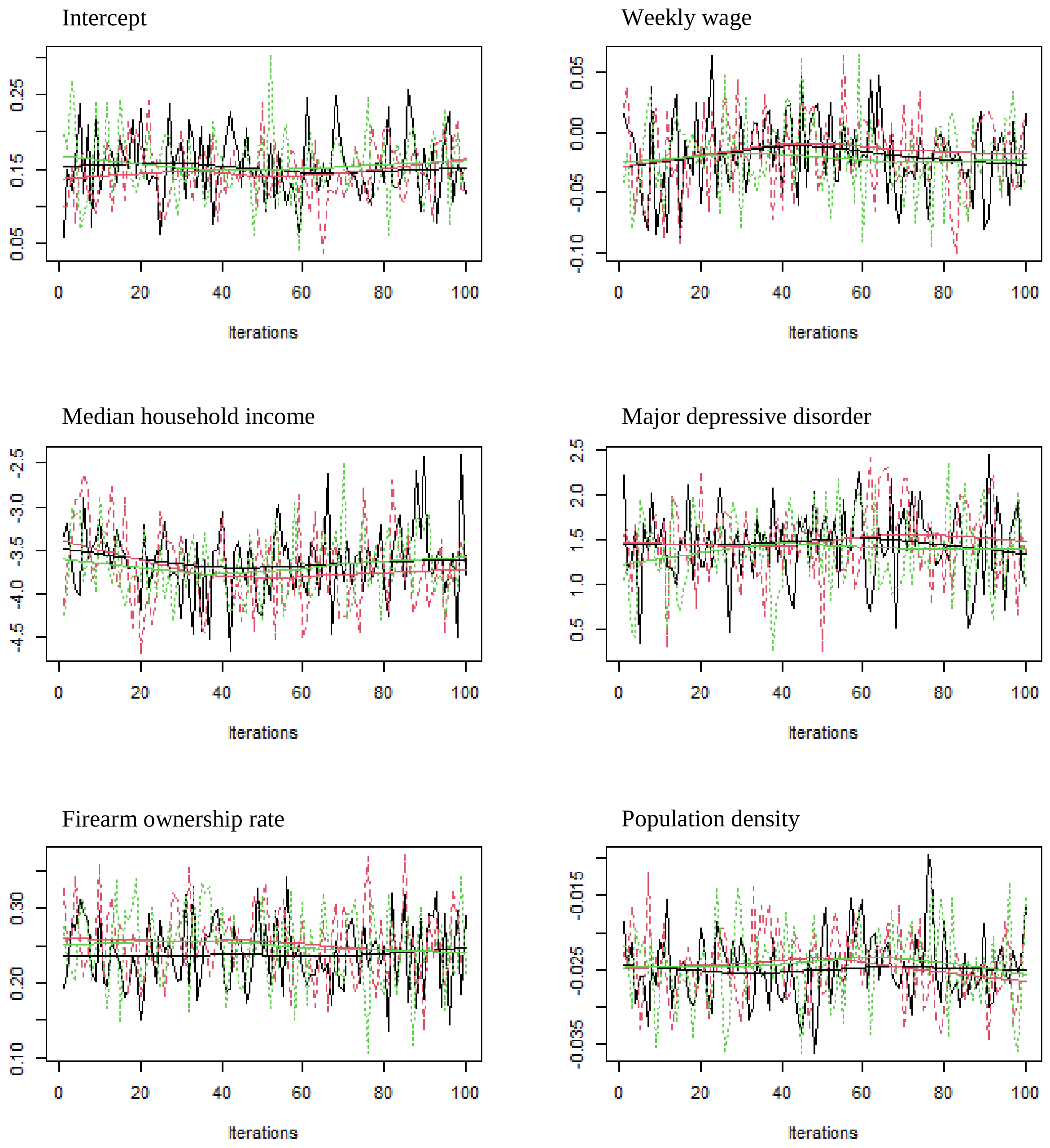


**Appendix Figure 4.** Trace plots for the intercept and five regression parameters in the *select* model over the fit period, for three chains (black, red, green), showing no clear trend in mean or variance, suggestive of chain convergence.

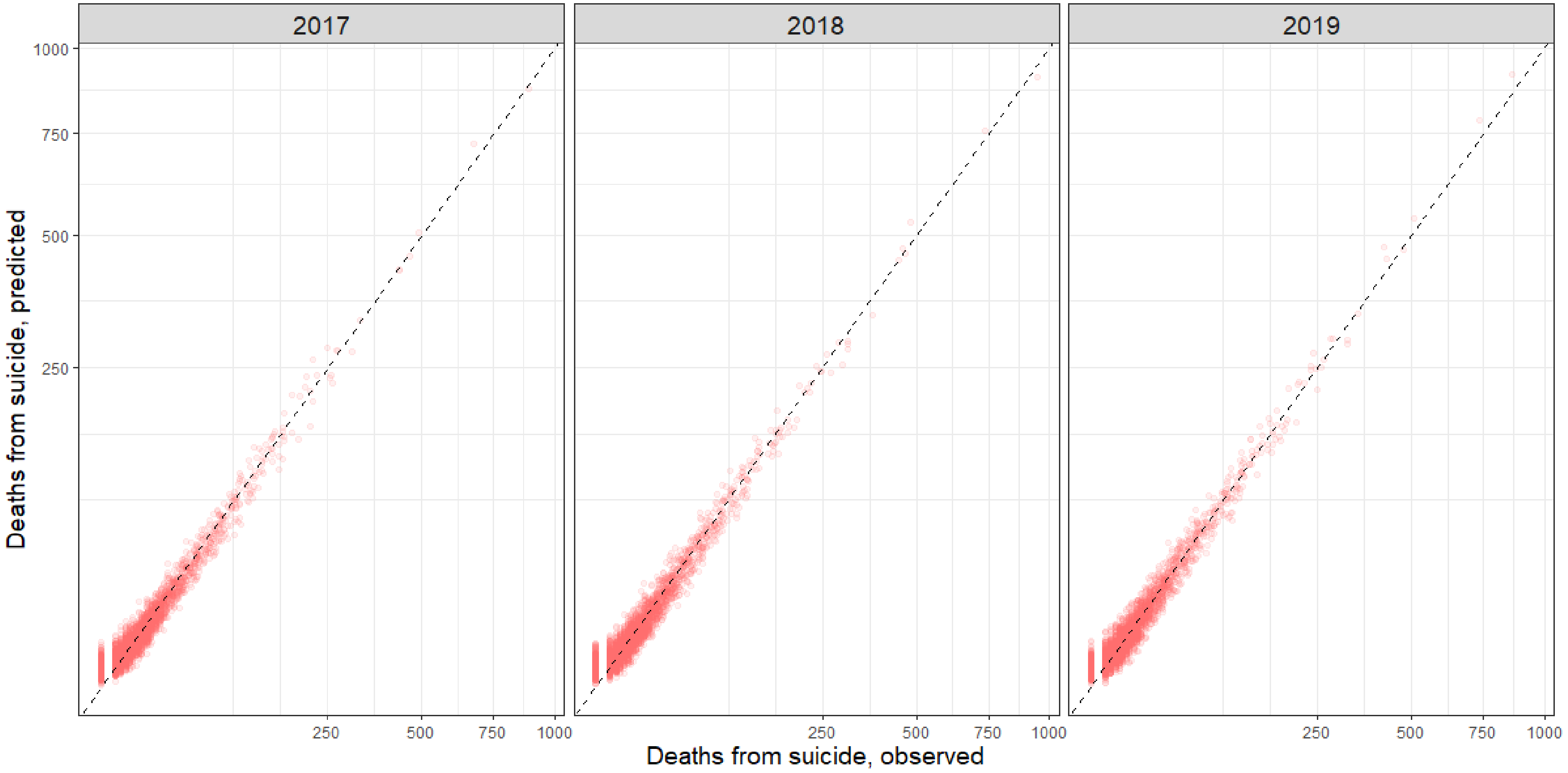


**Appendix Figure 5.** Scatter plot of model estimates of suicide deaths from *select* model in temporal OOS setting. Each data point represents a county. Axes are square root transformed.

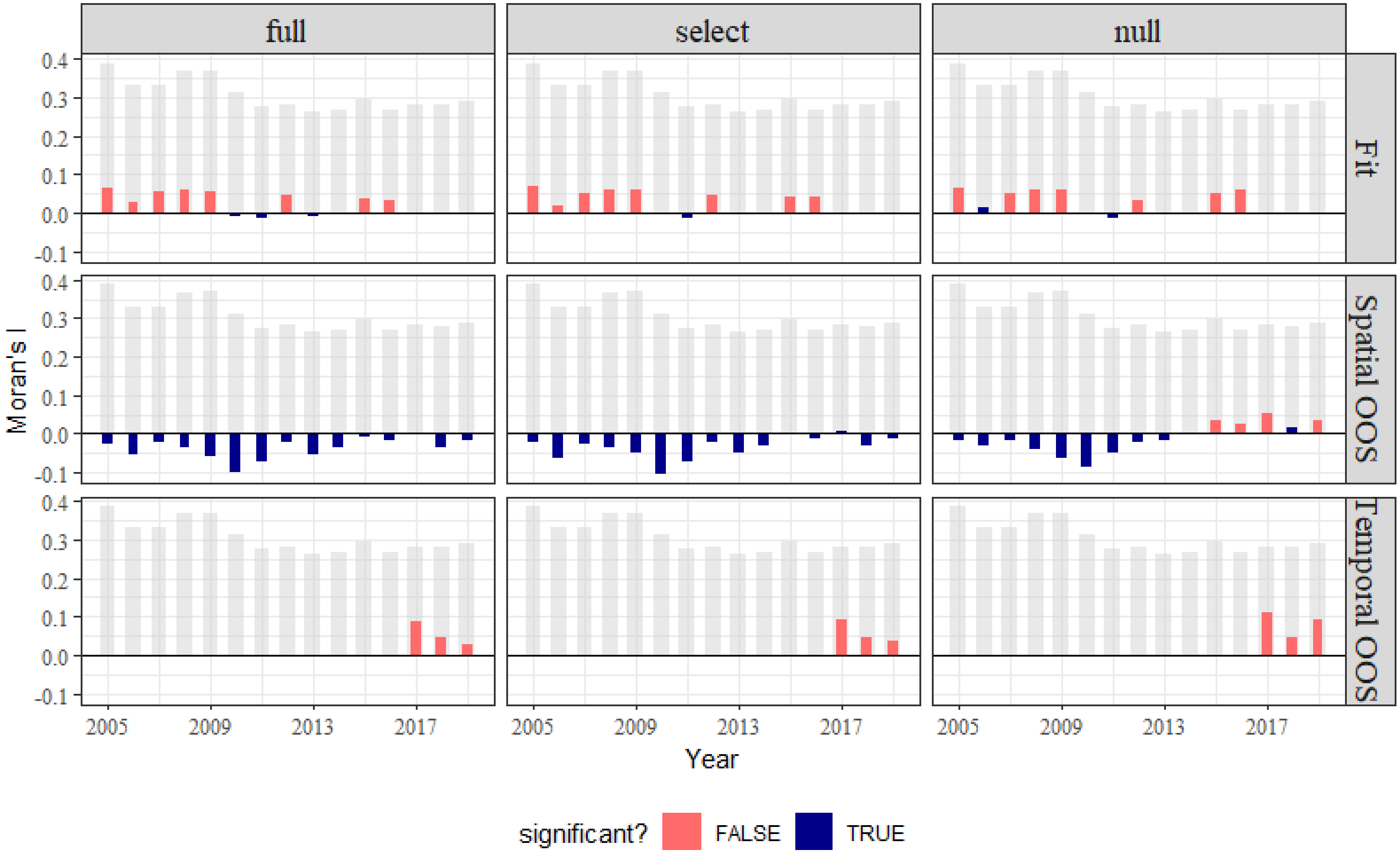


**Appendix Figure 6.** Global Moran' I in the residual. Magnitude of Moran's I statistic is indicated by the bars, with color indicating significance. The grey bars shows corresponding Moran's I from the *baseline* model (all significant).

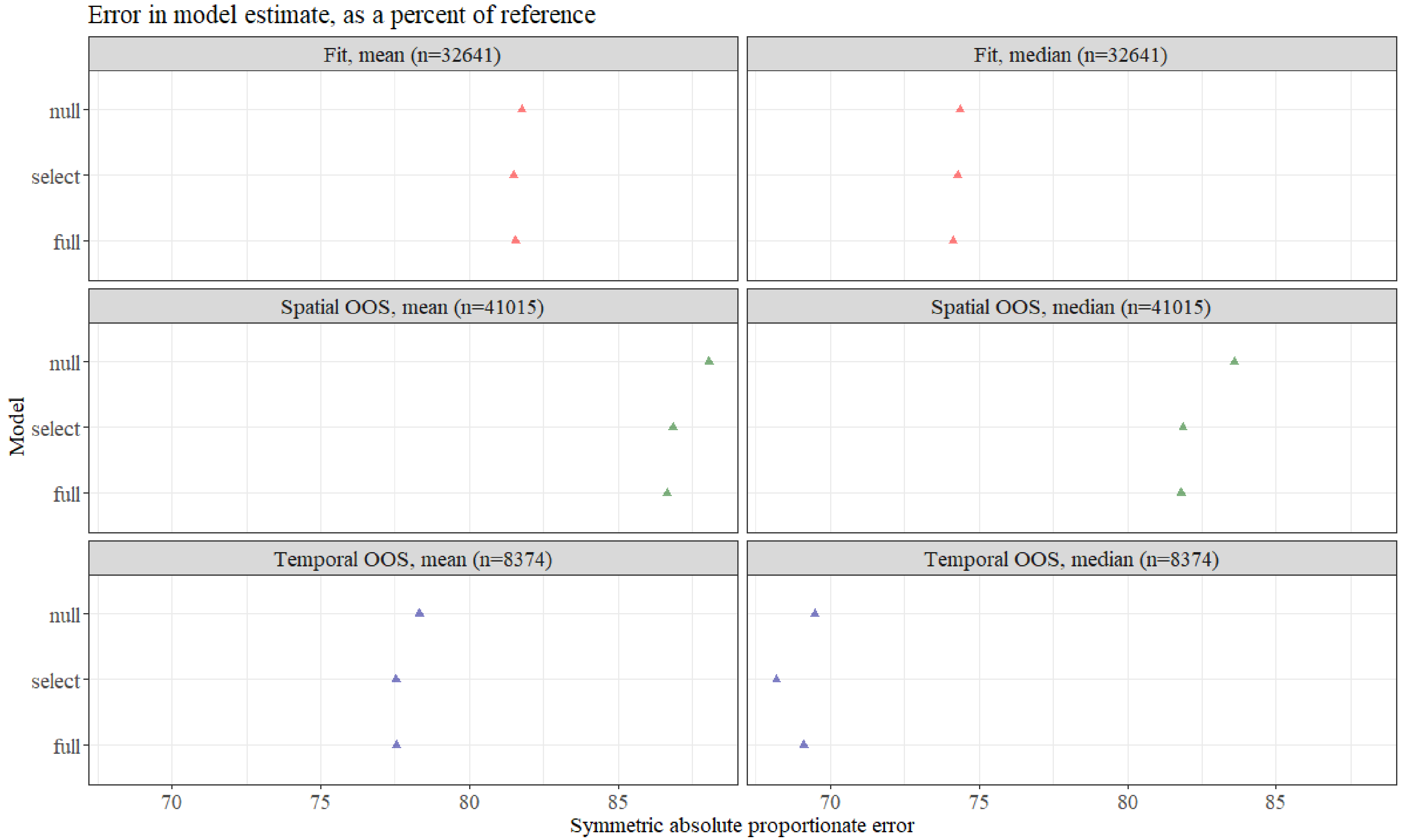


**Appendix Figure 7.** Mean and Median symmetric proportional error for in-sample, temporal out-of-sample and spatial out-of-sample estimates for all attempted model forms, as a percent of error in baseline estimates, *with zero-count county-year instances excluded*.

### References


A1. Gelman A, Little TC. Poststratification into many categories using hierarchical logistic regression. 1997.
